# Outdoor long-range transmission of COVID-19 and patient zero

**DOI:** 10.1101/2022.03.16.22272493

**Authors:** B.R. Rowe, J.B.A. Mitchell, A. Canosa, R. Draxler

## Abstract

Following the outdoor model of risk assessment developed in one of our previous studies, we demonstrate in the present work that long-range transport of infectious aerosols could initiate patient “zero” creation at distances downwind beyond one hundred kilometers. The very low probability of this outdoor transmission can be compensated by high numbers and densities of infected and susceptible people such as it occurs in large cities, respectively in the source and the target.

## I Introduction

Although it was originally discredited by governments and even health agencies, it is now well accepted that COVID-19 is mainly transmitted via aerosols (Wang *et al*., 2021). This has brought focus to the need for ventilation of interior spaces and the need for mask wearing, amongst general measures specific to this way of contamination. In a recent paper (Rowe *et al*., 2021), we have shown via simple modelling based on air flows, that the outdoor risk of being contaminated is generally several orders of magnitude less than indoors. Indeed, our paper uses concepts developed by Wells (Wells, 1955) and results in an outdoor model very close to the famous Wells-Riley model (Riley *et al*., 1978) for the probability of being infected by virions present in breathed air.

Another unknown in this epidemic is the infection’s origin. In the beginning, the question focused on the Chinese city of Wuhan, which has been the epicentre for the outbreak of COVID-19. Despite an extensive investigation, in particular focused on the Virus Research Laboratory or on a culprit animal that allowed the strain to jump over to the human host, no so-called “smoking gun” has emerged from these studies. Following this first onset, many countries then focused on the so-called “patient zero”, in order to prevent or circumvent an epidemic outbreak due to the cross border transit of a sick individual.

More recently we have seen the emergence of the more infectious Delta variant, apparently with an origin in India and sometime later, the even more infectious Omicron strain first identified in South Africa, but which quickly became rampant in the United Kingdom. Again we saw borders closing to try to contain this outbreak but with little success. As discussed in another of our recent papers (Rowe *et al*., 2022), the higher viral load or higher contagiousness of these new variants results in an even higher infectious risk by the aerosol route.

Based on our previous work (Rowe *et al*., 2021), what we investigate in the present paper, is the possibility that there is a long-distance airborne route for the passage of the virus from one region to another, leading to the creation of a few “patient zeros” who could serve as starting igniters of the epidemic in a new region. Of course, we insist on the fact that, in a given region, the epidemic itself cannot spread by this process alone, due to the extremely low probability of being infected in this way for a single individual (see Table 1). However, when the source of viral aerosol is a densely populated and extended area with a large infection prevalence and when the target itself is a territory of a fraction of a million of individuals or more, our model and calculations show that the emergence of patient zero is possible in this way.

**Table 1:**
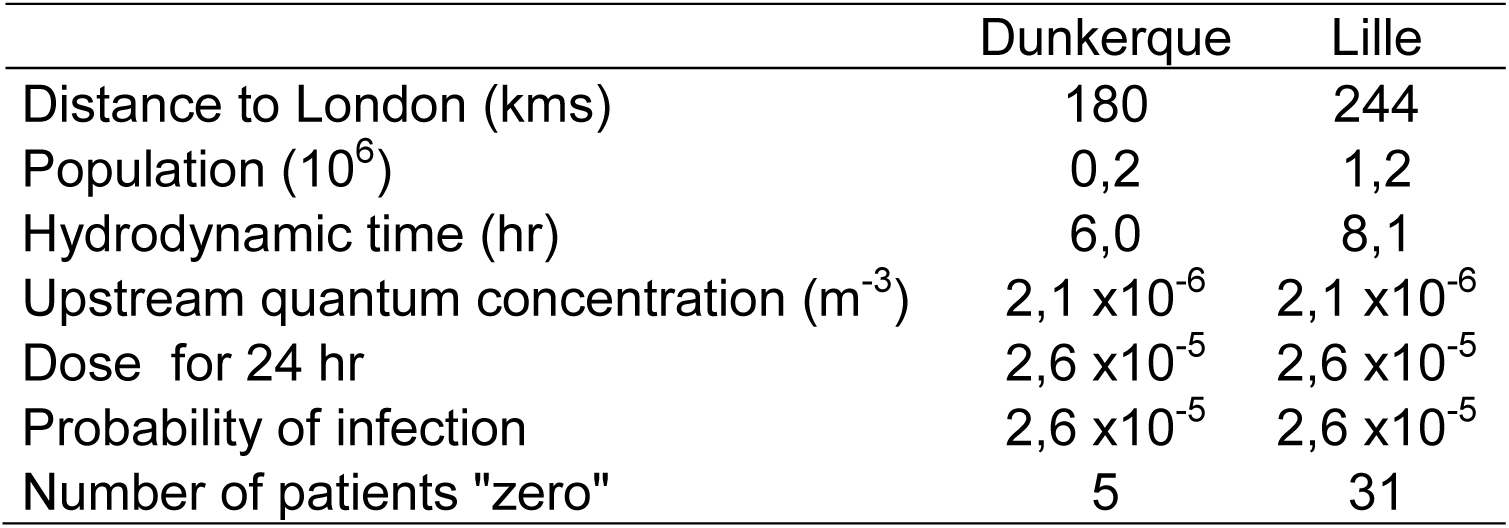
Possible number of “patient zeros” created by the long-distance transport of aerosols. London area population of 11 million; wind velocity: 30 km/h; exposure of 24 hours; proportion of possible infectors in Greater London: *r* = 3%; quantum production rate *q* = 10 h^-1^/infector.

|  | Dunkerque | Lille |
| --- | --- | --- |
| Distance to London (kms) | 180 | 244 |
| Population ($10^6$) | 0,2 | 1,2 |
| Hydrodynamic time (hr) | 6,0 | 8,1 |
| Upstream quantum concentration ($\text{m}^{-3}$) | $2,1 \times 10^{-6}$ | $2,1 \times 10^{-6}$ |
| Dose for 24 hr | $2,6 \times 10^{-5}$ | $2,6 \times 10^{-5}$ |
| Probability of infection | $2,6 \times 10^{-5}$ | $2,6 \times 10^{-5}$ |
| Number of patients "zero" | 5 | 31 |

The present paper is organized as follows: first we comment on some well-known cases of aerosol transport over very long distance, including infectious ones, then we establish our model of outdoor transmission in a slightly different manner than in our previous paper (Rowe *et al*., 2021), and in section IV we apply the model to a specific, although hypothetical, case: the possible contamination of Northern France by Southern England and London, especially with new variants of high viral load such as Delta or higher contagiousness as Omicron. Finally, we compare our atmospheric box model results to a 3-dimensional transport-dispersion model (HYSPLIT) calculation, discuss the lifetime virion question, and analyze our results in the light of the problem of contamination by very low dose, as related to single hit models (Teunis and Havelaar, 2000; Zwart *et al*., 2009; Haas *et al*., 2014; Brouwer *et al*., 2017; Rowe *et al*., 2022).

## II Long-distance transport of aerosols

The first thing to remember in the following discussion is that the SARS-CoV-2 virus is, first and foremost, a nanoparticle (diameter on the order of 100 nm). It is exhaled by an infected person (hereafter infector) in microdroplets of micron and submicron size (Greenhalgh *et al*., 2021; Tang *et al*., 2021). These microdroplets originate from respiratory fluids which, besides water as the main component, include a variety of other minor components: proteins, salt, etc. (Nicas *et al*., 2005). Water can evaporate leading to the creation of “dry nuclei” which include these minor components together with the virus. Due to the presence of non-volatile components, the reduction in size of the microdroplets cannot exceed a factor of around 0.4. Whether in aerosols or on surfaces, the virus is fragile. Its activity is influenced by temperature, humidity and ultraviolet (UV) light, which we will discuss further below. The fact that the virus is included in a microparticle means that it can float in the air for extended periods, driven by air currents both thermal and mechanical. The questions, therefore, are: can an infectious aerosol micro-particle travel over large distances? and if so, can it still have a biological effect after travelling? The second question centres around the survivability of viruses in an open environment and ultimately the statistics of the sources and receivers leading to a probability of “patient zero” creation.

The first question, concerning the long-distance transmission of micron and sub-micron sized particles, can be answered by experience and modelling. Examples of long-distance transmission of particulate matter in the atmosphere include the transport of Saharan sand (Francis *et al*., 2022), plastic microparticles (Allen *et al*., 2021), soot particles (PM_2.5_) from biomass burnings (Martins *et al*., 2018), and pollen transport from eastern North America to Greenland ((Rousseau *et al*., 2003) and references therein). These phenomena are well studied and documented and their importance evaluated. With the exception of pollen, these examples refer to non-biological, inert matter and are cited from the point of view of the coupling of observation and simulation to understand the modes and parameters associated with their transmission, and to demonstrate that long-distance travel can give rise to physical effects from these particles.

Does this long-distance transmission of microparticles have relevance to the spread of the COVID-19 virus? What is the likelihood of biological matter and in particular viruses inducing illness after long-distance transmission? In fact, these questions are now addressed by a part of science known as aerobiology, or aerovirology when restricted to viruses. In a recent review Dillon and Dillon (Dillon and Dillon, 2021) (and references therein) have pointed out the possibility of contamination on distances varying over several orders of magnitude and that long-distance atmospheric pathogen dispersion (500 m to 500 km) plays a crucial role in the propagation of a variety of plant and animal diseases.

There is much to learn from animal studies and indeed this is a transmission route that is taken very seriously by researchers worldwide for at least two cases which have considerable economic importance: Foot and Mouth Disease and Avian Flu respectively. The long distance airborne transmission of the first disease has been the subject of numerous publications ((Gloster *et al*., 1982; Donaldson *et al*., 1982; Hagerman *et al*., 2018), amongst others), including the possible transmission over the British channel between Brittany and Isle of Wight, i.e. for an oversea distance of around 300 km. Gloster et al. (Gloster *et al*., 2010) have presented the findings of a workshop held at the Institute for Animal health in the UK in 2008 that brought together researchers from the UK, US, Canada, Denmark, Australia and New Zealand to compare models for wind-borne transmission and infection of Foot and Mouth disease. Although the input parameters to the models (virus release, environmental fate, and subsequent infection) are undoubtedly sources of considerable uncertainty ((Gloster *et al*., 2010) and references therein), what was clear from this publication was that, under favourable meteorological conditions, the risk of long-distance infection was far from negligible. Nowadays the risk of windborne transmission of Foot and Mouth disease is still the subject of active research (Coffman *et al*., 2021) as this kind of transmission cannot be prevented by traditional epidemiological tools. Other studies have highlighted the long-distance transmission of the bird flu virus (Zhao *et al*., 2019) between farms in different states of the United States. It is significant that these studies take as a basis, that a single virus (or at least very few) can induce an infection (Sutmoller and Vose, 1997; Cannon and Garner, 1999).

Could a similar effect occur with the SARS-CoV-2 virus leading to outbreaks of COVID-19 without the necessity for a cross border transit of a sick individual? Let us look at statistics, probabilities and simple atmospheric models to see what they have to tell us.

## III Outdoor transmission: extension of a Wells-Riley type model

### III-1. Basic notions in airborne transmissions (indoor and outdoor)

Since the dawn of humanity, mankind has suffered of infectious diseases due to a variety of pathogens. In the recent decades, epidemiology has focused more on non-transmissible illnesses, such as heart disease, cancer, or obesity. However, the COVID-19 pandemic reminds us that the burden of infectious illnesses has not been eliminated.

Infectious diseases can be classified considering their target organs and the route of transmission following the path taken by the pathogen as it enters the body. As the target organs of the coronaviruses responsible of the COVID-19 are located in the respiratory tract, this disease can be classified as respiratory. As discussed in (Rowe *et al*., 2021) and references therein, the route of transmission has been a matter of intense debate but, as stated in the introduction, it is nowadays largely recognized that the major transmission path is through airborne exchange i.e., by inhalation of an aerosol that has been exhaled by an infected person.

In matters of infectious disease and epidemiology, a key problem is to assess the dose-response relationship i.e., what is the probability of infection resulting from a given level of exposure for a given time (the dose) to a pathogen (Brouwer *et al*., 2017). The dose *X* is clearly linked to a number of pathogens. A dose-response function *P*(*X*) relates the dose to a probability of infection. It is clear that *P*(*X*) must be a monotonically increasing function of the dose, starting from zero at zero dose and increasing toward an asymptote *P* = 1 for large values of *X*. There are several probability laws that can be used for *P*(*X*) as discussed in (Brouwer *et al*., 2017), one of the most widely used being the exponential form:

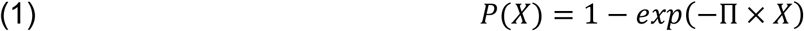

where Π is a numerical factor which depends on the choice of the dose counting unit reference. In fact, one of the recognized difficulties in the dose-response model is first to define the dose. It is beyond the scope of the present paper to examine this question in detail and the reader is referred to the book of Haas (Haas *et al*., 2014) and to (Brouwer *et al*., 2017). For an airborne disease, **Wells** (Wells, 1955), using the exponential law, **defined a quantum of contagium** as a hypothetical quantity that has been inhaled per susceptible individual when 63.2% (corresponding to 1 − exp(−1)) of these individuals display symptoms of infection. It is linked to a probability of infection which then follows a Poisson law:

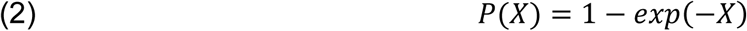

The quantum has no dimension **but is a counting unit** (as dozens versus unity, or moles compared to molecules) which is clearly linked to a choice of Π = 1 in eq. (1). Of course, and as discussed in (Brouwer *et al*., 2017) and (Rowe *et al*., 2022), its value, in terms of the number of pathogens, depends on a variety of mechanisms: inhalation of airborne particles, pathogen inhibition by host defenses or losses by some other processes, before any replication will start in an infected cell. Obviously, a quantum corresponds statistically to a number of pathogens much greater than one.

Considering the concentration of quanta in space (in m^-3^ units), 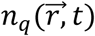 the inhaled dose during a time of exposure t can be written as:

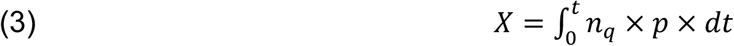

*p* being the pulmonary ventilation rate (taken as 0.5 m^3^/h in the present investigation). Note that this definition of the dose does not require a homogeneous distribution of quanta in space. Only 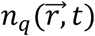 at the inhaled location (mouth and nostrils) has to be considered. Note also that due to the extremely low concentration of quanta in air, 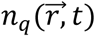 is not really a continuous function of 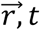 (since a number of viruses is of course an integer) but can be treated as such due to the statistical aspect of the solution.

As shown in our previous papers (Rowe *et al*., 2021; Rowe *et al*., 2022), in the case of an indoor room with well-mixed air, it is possible to write a conservation equation for the quanta, which, together with eq. (3), leads directly to the stationary state dose value and to the well-known Wells-Riley probability:

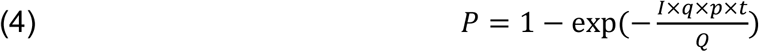

where *P* is the probability of infection for a susceptible person, *q* a quantum production rate per infector per unit time, *p* the pulmonary ventilation rate, *Q* the ventilation rate of the room, *I* the number of infectors in the room, and *t* the time of exposure.

### III-2. Box model of outdoor transmission

As discussed in the previous section, the first step in any models, indoor or outdoor, is to evaluate the concentration of the virions (which can be counted in terms of quantum) in inhaled air. The outdoor model developed in one of our previous papers (Rowe *et al*., 2021) is essentially a “box” model as described in chapter 5 of Environmental Impact Assessment (Mareddy, 2017), and developed previously by numerous researchers (Nelson and LaBelle, 1975; Ortolano, 1985; Canter, 1986). Box models are based on mass balance equations and are the simplest atmospheric models that can be used to evaluate the mean concentration of pollutants (molecules or particles), downwind of a source.

Our 2021 models considered mono-sized infectious microdroplets and their airborne behavior. In the following paragraphs, we develop, in some detail, an identical outdoor box model, using the Wells notion of quantum for the counting of virions. We consider an outdoor volume (atmospheric box) as illustrated in figure 1, with the wind blowing along the x axis and, as in our previous work, we suppose that there are no quanta escaping the volume above a height *H* along z. The evaluation of *H* is the most critical part of the model. It is also assumed that the quantum density does not change across the wind:

**Figure 1:**
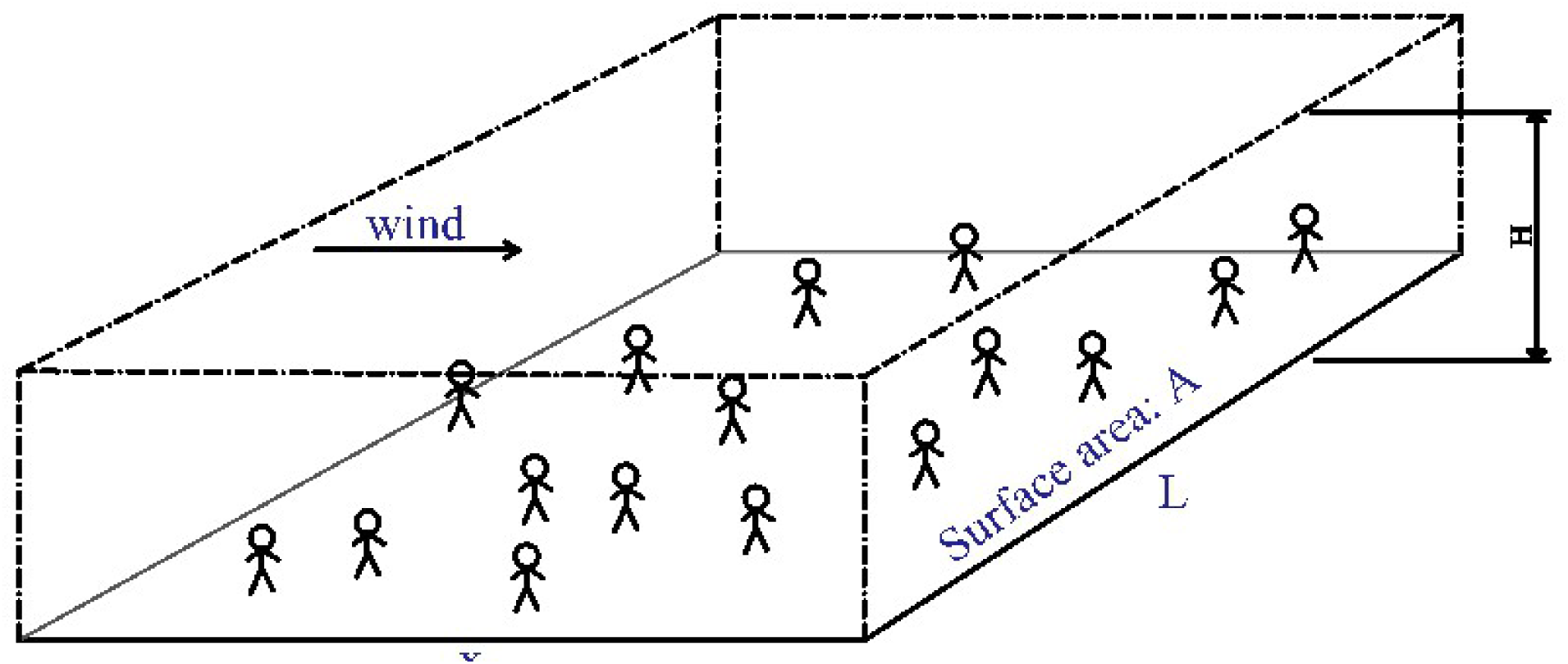
The atmospheric box model

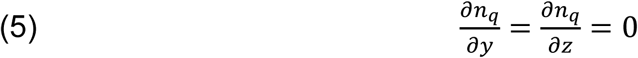

and hence the quantum concentration is considered only as a function of *x* along the wind. Although at low values of *x, n*_*q*_ is a function of height *z*, assuming that eq. (5) holds everywhere, the height dependency does not change the concentration balance between what is produced in the bulk of the box and what emerges at its downwind border at large *x*, where everything has been mixed by the turbulent dispersion. This assumption is inherent to box models (Ortolano, 1985). It can, therefore, safely be concluded that eq. (5) has no influence on the quantum concentration at this border.

Then, assuming stationary state i.e., 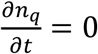, a conservation equation for the quanta can be written as:

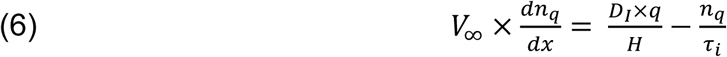

where *D*_*I*_ is a density of infectors per unit surface (assumed homogeneous and therefore constant), *V*_∞_ is the wind velocity and *τ*_*i*_ is the virus lifetime defined from the temporal exponential decay of active virions in microparticles, due to natural physicochemical processes. Note that with this definition, *τ*_*i*_ is slightly different from the so-called half-life which is the time required to decrease the active virion concentration by a factor of two (since here *τ*_*i*_ corresponds to *exp*(−1) = 0.37).

Note that the infectors are located at the bottom of the atmospheric box (which can include houses as we will discuss in section IV-3) but this has no influence on the calculation since we assumed an homogeneous dispersion of the viral aerosol in the vertical dimension of the box, as discussed in previous paragraph.

With a quantum concentration *n*_*q*_(0) at *x* = 0, we can derive the following value for the quantum concentration:

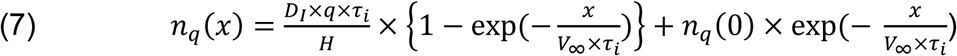

In an area where there is no infector, *D*_*I*_ = 0, eq. (7) leads to a simple downwind exponential decay of the quantum concentration:

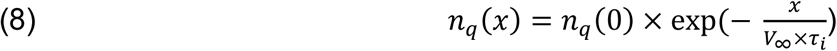

On the other hand, and for a virus lifetime much longer than the hydrodynamic time *τ*_*h*_ (i.e., *τ*_*i*_ ≫ *τ*_*h*_ = *e*/*V*_∞_), eq. (7) leads to the following value for *n*_*q*_(*x*):

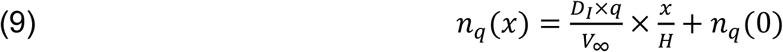

This is analogous to the equation derived in (Rowe *et al*., 2021) for *n*_*q*_(0) = 0, and which expresses the conservation of quanta in the atmospheric box shown in figure 1 when there is no decay due to viral inactivation.

Using equation (2) and (3), it is then possible to calculate the probability of infection at a distance *x*. In equations (7) and (9), **a key parameter is the value of *H* and therefore** 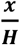, as discussed at length by Rowe et al. (Rowe *et al*., 2021) in their supplementary materials. For strong winds i.e. *V*_∞_ > 6m/s at 10 m height and night time or low solar insolation, the atmosphere can be considered as neutral in the so-called Pasquill– Gifford– Turner classification (Pasquill, 1961; Gifford, 1961; Turner, 1994), which means there is no tendency for turbulence in the air to be enhanced (unstable) or suppressed (stable) through the buoyancy effects. In fact, it is admitted that the airborne pollutants emitted locally are transported and dispersed within the so-called atmospheric boundary layer (ABL: the tropospheric bottom layer), whose thickness is usually lower than one thousand meters (Sáez de Cámara Oleaga, 2016), excepted for strongly unstable atmospheres. At any distance from the source, an order of magnitude value of *H* versus *x* can be estimated by the vertical dispersion length used for Gaussian plumes and shown in figure 2 (Turner, 1970; Seinfeld and Pandis, 2016).

**Figure 2:**
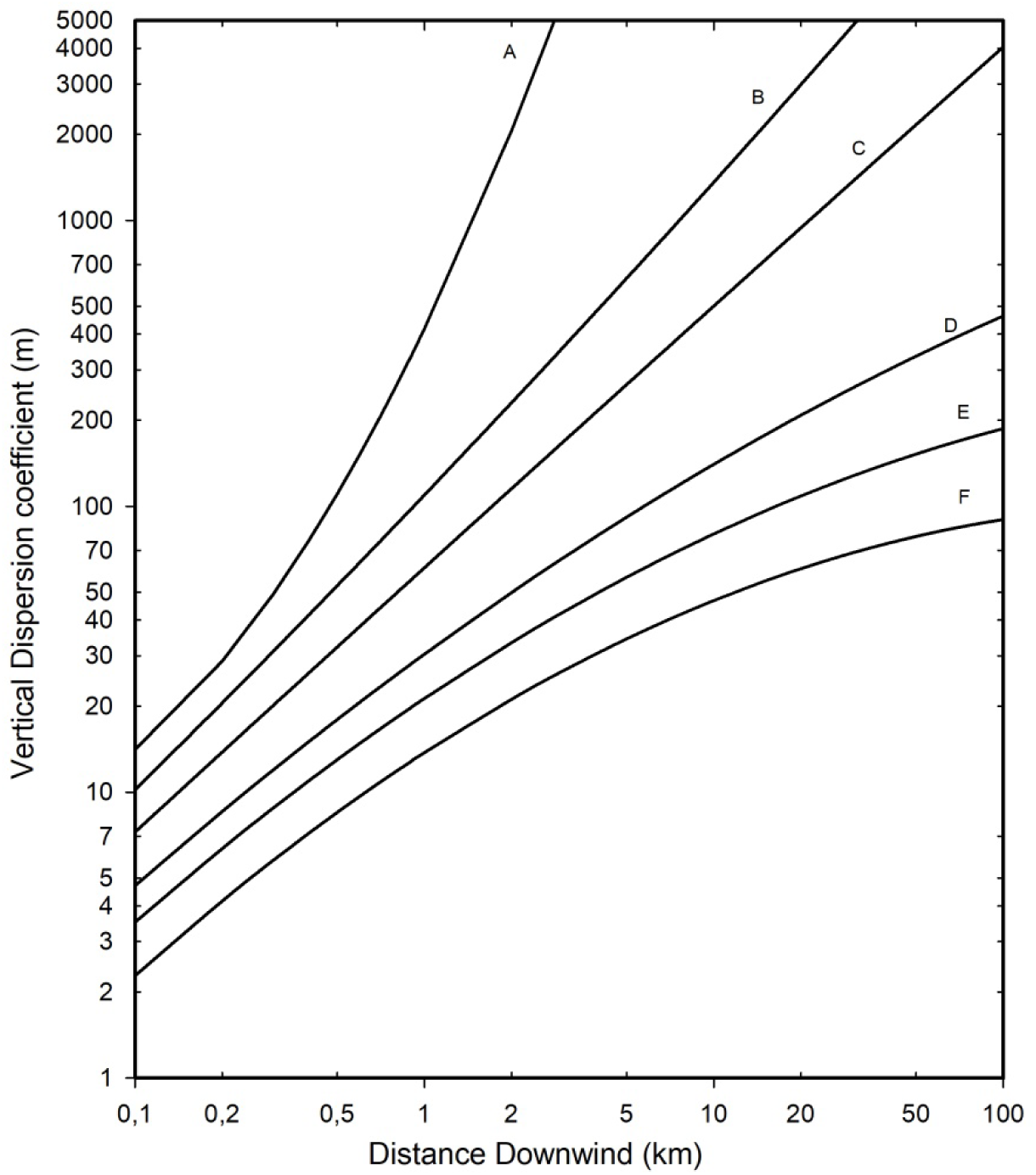
vertical dispersion length for Gaussian plumes. Classification of atmosphere state: A: Extremely unstable; B: Moderately unstable; C: slightly unstable; D: neutral; E: slightly stable; F: moderately stable.

The areal density of infectors can be taken as:

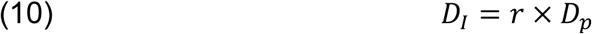

with *r* the proportion of infectors and *D*_*p*_ the areal density of population in the space. The condition 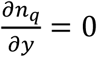 requires that there is no gradient in mean infector density across the wind. If we assume a value of *L* for the width of the source, then, for *n*_*q*_(0) = 0 eq. (9) also reads:

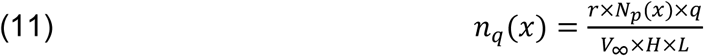

where *N*_*p*_(*x*) = *D*_*p*_ × *x* × *L* is now the total population in the area *x* × *L*.

The wind itself depends on the altitude but its variations above ten meters are rather small within the ABL (Hsu *et al*., 1994). Therefore, in the next section it will be assumed to be independent of altitude and taken as the ten-meter value.

## IV Possible airborne creation of “patient zeros” by long-range transmission

### IV-1. General considerations

Imagine two strongly populated areas, designated as the “source” and the “target” respectively, separated by an unpopulated area (no man’s land). At the downwind border of the source, it is possible to quantify the quantum concentration following the above equations. Its evolution in the “no man’s land” will be ruled only by the virus lifetime following eq. (6) with *D*_*I*_ = 0. This will lead to its new value at the upstream border of the target area. Assuming no evolution of the quantum concentration downwind (consistent with the virus lifetime discussion of section V-2) or across the wind (which will be the case if the width of the target is smaller than the width of the source) then the calculation of a probability of infection *P*_*t*_ in the target area of population *N*_*t*_ is straightforward.

It follows that the statistical number of contaminated susceptible people *S*_*c*_ is:

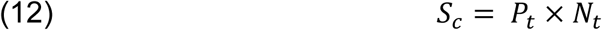

As will be shown further, the value of *P*_*t*_ will most often be extremely small, which shows a quasi-zero risk at the individual level. However, if the target is composed of a very high number *N*_*t*_ of individuals, then a few people (≥ 1) could be infected. Of course, this process alone cannot sustain an epidemic but creates a few infectors (“patient zero”) which will ignite it. Note that *N*_*t*_ is the total number of individuals in the target, assumed healthy, and therefore susceptible.

In winter and early spring, the strongest winds are most often from west to east in Western Europe. Below we examine a hypothetical case study of the creation of patients zero in Northern France from Southern England in wintertime.

### IV-3. Model of long-range transmission from Southern England to Northern France

An infographic image of a model of three boxes is shown in figure 3. Following the discussion of the previous section, we consider the first box in Southern England corresponding to the upstream source of contamination. Grossly, it corresponds to the Greater London area and its downwind region. Although the county of Kent, in the southeast of London, is quite heavily populated, we shall restrict ourselves to a source around London with a width *L* of 40 kilometers and a length *x* of 45 kilometers where we assume a population of 11 million people.

**Figure 3.**
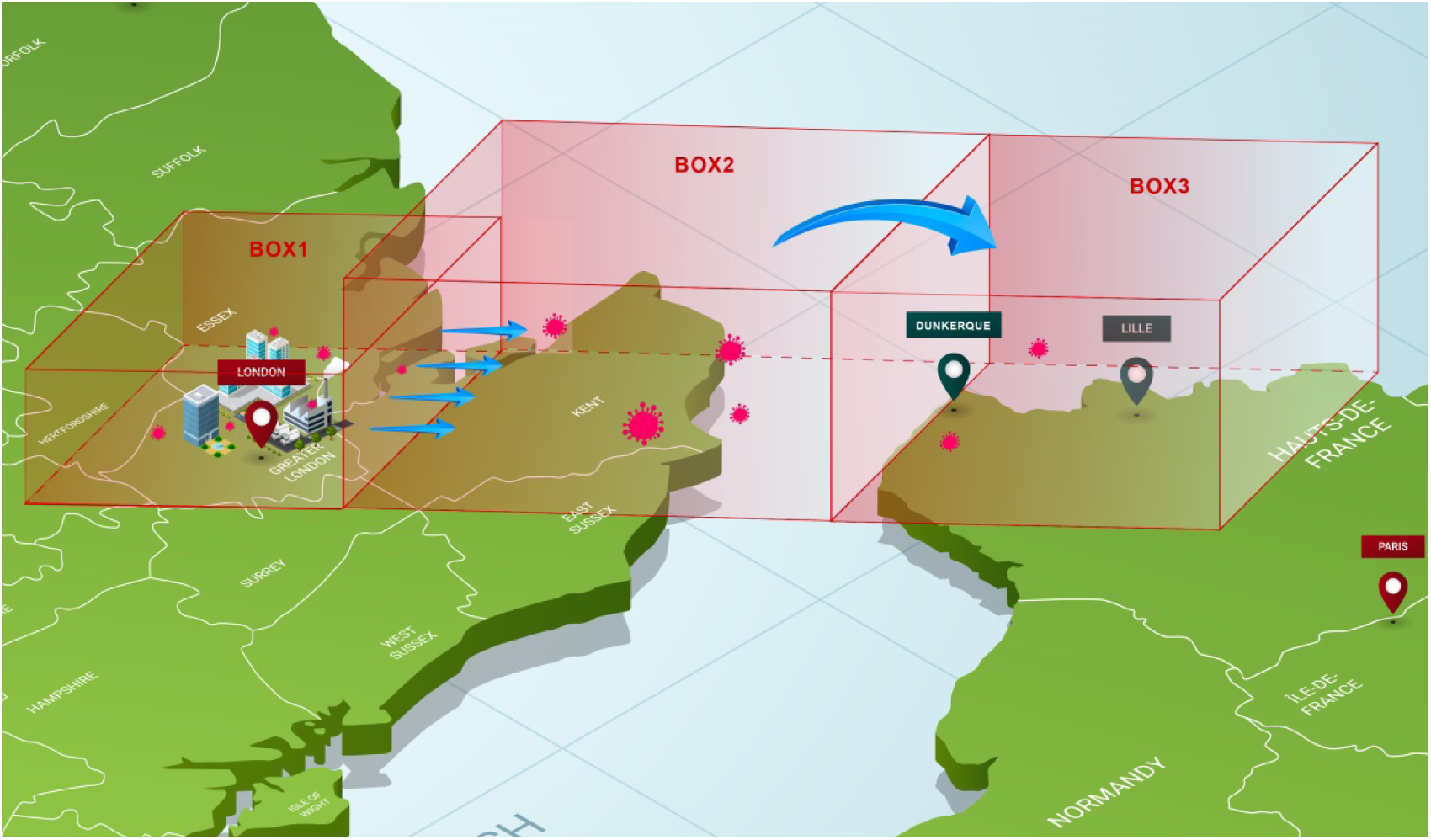
model of three boxes between Greater London and Northern France

We can use the equations developed in section III-2 to make an estimate of the quantum concentration at the downwind border of this source box. However, the following problem arises: in wintertime most of the quanta will be emitted indoors, with a room temperature around 20 °C, and a rather low RH (we assume 35 % as a mean), but outdoors they are transported by the wind at low temperature (around 5°C) and rather high humidity (80%) conditions, where the virus lifetime is expected to be much longer than the atmospheric transport (hydrodynamic) time. Therefore, viral inactivation, as discussed in section V-2, will only occur indoors, via thermal effects at rather low RH. Indoor air is continuously renewed as contaminated air is exhausted outdoors with a characteristic time equal to 1/*ACH* where *ACH* is the air change per hour. Therefore, the effect of viral inactivation indoors prior to exhaust can be taken as a reduction of the quantum emission rate per infector used in section III-2 following the formula:

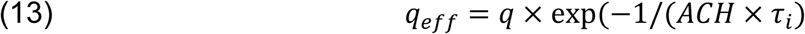

with the conservative hypothesis of *ACH* = 2 *h*^−1^ and *τ*_*i*_ = 2 *h* (which is a good order of magnitude at 20°C and low RH, see references of section V-2), this results in *q*_*eff*_ = 0.78 × *q*. Note again that *τ*_*i*_ relates here to indoor conditions at the source.

Once outdoors, the typical wintertime low temperatures will ensure a very long virus lifetime. Thus, the simple formula (11) can be used, with *q*_*eff*_ instead of *q*, to estimate the quantum concentration at the downwind border of the source. *H* is estimated from figure 2 as 300 m (for *x* = 45 km and state D in figure 2), the population within the source as 1.1 × 10^7^, the wind velocity taken as 30 km/h, the width of the source as *L* = 40 km (which influences the density of infectors if formula (9) is used in place of 11)), and the quantum production rate of an infector as 10 h^-1^. The numerical application leads to *n*_*q*_(45 *km*) = 7.15 × 10^−6^ *m*^−3^ assuming a proportion of infected persons of r = 0.03 in the Greater London area.

Turbulent mixing and transport by the wind then will lead quanta at the upstream border of the target area whose width is assumed less than or equal to the width of the source. Again, due to meteorological conditions (mean temperature around 5°C, mean RH around 80% (Weather and Climate, 2022), and absence of UV radiation), the virus lifetime is much higher than the convective hydrodynamic time (which is <10 h, see Table 1) for distances up to 250 km and therefore the reduction in quantum concentration is solely due to the increase of the dispersive height *H*. Using a conservative estimate for *H* of 1000 m, a value corresponding to a common upper value of ABL thickness for neutral or stable conditions (Sáez de Cámara Oleaga, 2016), results a numerical value of *n*_*q*_ of 2.15 × 10^−6^ *m*^−3^ at the upstream border of one of our targets.

We consider two plausible targets in France, either the city of Dunkerque or the Lille agglomeration, with populations of 0.2 × 10^6^ and 1.2 × 10^6^ respectively. We assume that the wind direction is the same as the direct path between the source and the target, a dominant direction in wintertime, which grossly corresponds to a wind direction from the west/northwest (respectively 288 and 294 degree). As before, we also assume a wind velocity of 30 km/h which is only slightly higher than the mean wind velocity in February/early March (Weather Sparks, 2022). Note again that both target areas have a width across the wind less than that of the source. Table 1 summarizes the assumed values of various parameters leading to a statistical number of infected “patient zeros”. Since this number appears to be a few units in the frame of our assumptions, it clearly reveals the potential possibility of an infection ignition through long-range transportation of airborne viruses.

Note that the purpose of the calculations presented in Table 1 is solely to demonstrate the possible creation of patient “zero”. Changing the wind speed and other parameter values in a reasonable manner does not alter this conclusion. Also, it could be possible to refine the calculation considering a much longer time of exposure (several days) and the fact that virions in the target box can be inactivated indoor by thermal and RH effects like assumed for the source box, but again without changing the major conclusion.

From this table it can be inferred that creation of only one patient zero is possible within a day even with a much lower proportion of infectors (*r* ∼ 0.3%).

## V Discussion

### V-1. Validity of the atmospheric box model

To determine if the box model assumptions were sufficiently realistic, a test calculation was performed using a three-dimensional particle transport and dispersion model. The HYSPLIT model (Stein *et al*., 2015) was selected for the simulation using one-degree resolution gridded global meteorological data available at three-hour intervals from the National Oceanic and Atmospheric Administration (NOAA). As noted earlier, west to east flow is common, and a 24 h period (starting 0600 UTC 5 January 2022) with airflow from London to France was identified in the first week of data downloaded from the GDAS server (National Centers for Environmental Information, 2022). The model was configured to be similar to the box model. A 40 km line of five-point sources, orthogonal to the wind direction, was set over central London with a total emission rate of one unit per hour for the 24 hour simulation period. Air concentrations (unit mass over volume) were computed as a 24-hour average on a 10 km resolution grid with a vertical depth of 100 m to represent the surface layer for human exposure. Particles were terminated after 12 hours to constrain the downwind domain to France rather than all of Western Europe. The HYSPLIT result in Fig. 4 shows a broad region of concentrations of around 5×10^−14^ m^-3^ over northeastern France. Using the effective quantum production rate *q*_*eff*_ value derived from Table 1, the unit emission HYSPLIT values, through the expression: *N*_*P*_×*r*×*q*_*eff*_×24, convert to a quantum concentration of 3.1×10^−6^ quantum m^-3^, which is very close to the upstream box model value of 2.1×10^−6^. Due to the line source configuration, lateral dispersion along the centerline would be negligible and the concentration results would primarily depend upon the vertical mixing. An examination of the diagnostic vertical mass profile after 12 hours (not shown) indicates that 94% of the mass was in the first 1200 m above ground and 99% was within the first 1500 m, consistent with the well-mixed box model assumptions.

**Figure 4:**
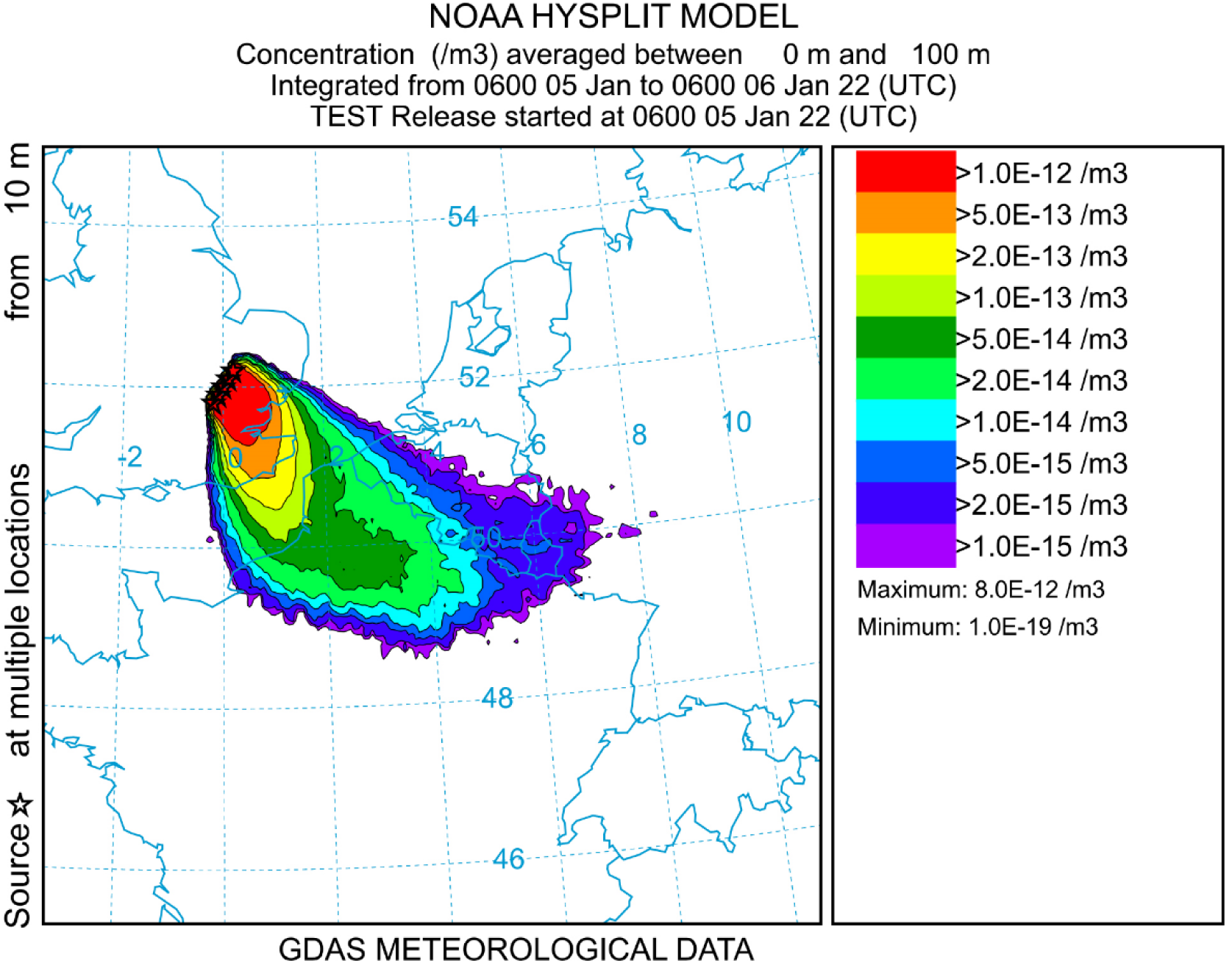
results of the HYSPLIT model

### V-2. The question of the virus lifetime in aerosol form

The virus lifetime used in the above atmospheric model is defined, as stated above, by the analog in time of eq. (8):

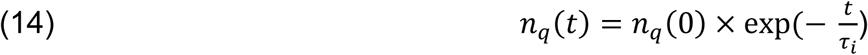

As shown in the previous sections, the effect of virus lifetime is critical for the possibility of virus transmission over long distances. If, for *τ*_*i*_ ≫ *τ*_*h*_, the decrease of quantum concentration downstream of a laterally extended source is mainly due to vertical atmospheric dispersion, for *τ*_*i*_ ≪ *τ*_*h*_ the quantum concentration will drop to an even much lower value, with a ratio of *τ*_*i*_/*τ*_*h*_. This condition corresponds to a distance to the source *x* ≫ *τ*_*i*_ × *V*_∞_ where the transmission probability drops essentially to zero.

Viruses are inactivated by a variety of factors, but it is recognized that the principal ones are temperature, humidity, and UV radiation. Although solar UV radiation is very efficient for virus inactivation (Lytle and Sagripanti, 2005), we shall not consider it here, restricting ourselves to the case of mean and high latitudes in winter, where nighttime is much longer than daytime, and where the sky is often overcast during the day, making UV inactivation negligible outdoors during atmospheric virus transport. In fact this conclusion holds even for a clear sky, at least from early November to the end of February, at a latitude around 51°N due to large solar zenith angle which then reduces by orders of magnitude the efficiency of UV virus inactivation, due to ozone absorption of solar UV (Lytle and Sagripanti, 2005).

The effect of humidity on lifetime is rather difficult to assess (Ijaz *et al*., 1985; Yang and Marr, 2012). After some discussion (Marr *et al*., 2019) about whether to consider absolute humidity (AH) or relative humidity (RH), it is concluded that RH drives the virus lifetime with a U-shaped curve for *τ*_*i*_ = *f*(*RH*). This behavior is due to the variation of the solute/solvent concentrations in the aqueous solution of the infectious microdroplet. This U-shape has been rationalized by Morris et al. (Morris *et al*., 2021) considering the efflorescence relative humidity (ERH) which corresponds to the RH below which a spontaneous evaporation of a salty water solution initiates a crystallization called efflorescence (leading then to what is called dry nuclei for viral aerosols). Note that the inverse process, when RH is increased, is called deliquescence (Horst *et al*., 2019).

The inverse of the lifetime *k* can be considered as a rate of inactivation (unit = time^-1^) and it is generally admitted (Yap *et al*., 2020; Morris *et al*., 2021) that, for a given value of RH, it follows an Arrhenius law with temperature:

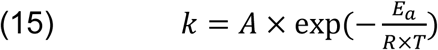

On the basis of numerous experimental results for a variety of viruses, this behavior has been rationalized by Yap et al. (Yap *et al*., 2020) as viral protein denaturation by thermal effects, including for SARS-CoV-2. In this last case, Yap et al. report values of *E*_*a*_ = 135.69 kJ/mole and *A* = 1.3×10^21^ min^-1^ respectively.

There are clearly large uncertainties in the exact lifetime of SARS-CoV-2 in aerosol form at given values of RH and temperature. For example, van Doremalen et al. (van Doremalen *et al*., 2020) report a value of 1-3 hours at a temperature of 21-23 °C and RH of 40% although Fears et al. (Fears *et al*., 2020) found a much higher value (up to sixteen hours). At the opposite, a group from Bristol (UK) has recently reported a very fast loss of infectivity (seconds to minutes), of virions aerosolized from a tissue culture medium, hereafter TCM, at room temperature (Oswin *et al*., 2022). Note however that Oswin *et al*. do not observe an exponential decrease, instead a plateau after an initial rapid loss, preventing the determination of a virus lifetime as defined by eq. (14). Note also that (Smither *et al*., 2020) have reported very different losses of infectivity between aerosols generated from TCM and artificial saliva and that, in the real life, respiratory fluids should be used. A short (few minutes) lifetime in aerosols for normal indoor conditions is not compatible with airborne transmission which has been largely documented (Wang *et al*., 2021).

The large uncertainty in lifetime can be understood due to the extreme difficulty of virus concentration measurements in air and, worse, of their characterization as to whether they are active or not (Haas *et al*., 2014).

In any cases, based on most of the above studies and others (Haas *et al*., 2014; van Doremalen *et al*., 2020; Yap *et al*., 2020; Morris *et al*., 2021), the order of magnitude of the virus lifetime as defined by eq. (14), is around one to a few hours at room temperature. It can then be easily shown that at low temperatures (<5°C) and large RH (around 80%), SARS-CoV-2 has a lifetime at least of several tens of hours, an important conclusion which has been used in our box model.

### V-3. The very low dose question

That a single susceptible person inhales numerous virions transported by the wind over long distance is of course highly unlikely. Therefore, in the framework of the present paper, the question of the dose/risk function at very low dose deserves a careful discussion.

The literature on virus transmission very often refers to a quantity named “Minimum Infective Dose” with the acronym MID. But, as stated by Haas et al. (Haas *et al*., 2014) the term “Minimum” is very misleading as it seems to imply a minimum number of pathogens needed to start an infection and should be replaced by “Median”. A real minimum would imply a thresholding effect which is not observed experimentally. If such an effect existed, it would prevent the long-distance transmission presented above. However, there exists a very large literature on the subject of the dose/risk link (Teunis and Havelaar, 2000; Zwart *et al*., 2009; Haas *et al*., 2014; Brouwer *et al*., 2017) and most authors conclude that a single pathogen can trigger infection (Brouwer *et al*., 2017), although with a small probability (single hit models). The exponential dose-risk function used in the present paper is clearly without threshold and used widely elsewhere. Together with the quantum concept, it completely takes care of the statistical and probabilistic aspects of the transmission problem.

## V Conclusion

In the present paper we have shown, with a rather simple model of conservation equations and a dose-risk function, that creation of “patient zero” at large distance from a densely populated and infected area is possible if the target is a large population. Surprisingly, this is well-known and accepted in veterinary science but the link with human airborne transmission, to our knowledge, has not been made. Our simple atmospheric “box” model has been validated by a 3-dimensional dispersion (HYSPLIT) calculation, using actual meteorological data. One of the consequences is that the search for “patient zero” could sometimes be meaningless. It also shows that, at the level of a continent, viruses ignore borders and that there is no need of personal travel, to spread infection downwind of a contaminated region. However, due to the importance of climate on the virus lifetime, it has to be kept in mind that the conclusions and hypotheses presented here **apply mainly to mid and high latitudes under winter conditions**.

Although it has not been discussed in the present paper, note that in February 2021 most of the contamination in Dunkerque was due to British variant (France Info, 2021) which could have started by the process described above. It would be of course extremely interesting to conduct a more refined analysis than the one presented here concerning a longer period of time and using available epidemiological and atmospheric data. The purpose of the present study was only to highlight the possibility of the process.

## Data Availability

All data produced in the present work are contained in the manuscript

## Acknowledgments

Bertrand Rowe wishes to thank Dr. Daniel Preston, of Rice University, for very helpful discussions on virus lifetimes and Melinda Rowe for her help as digital designer.

